# Serial interval and basic reproduction number of SARS-CoV-2 Omicron variant in South Korea

**DOI:** 10.1101/2021.12.25.21268301

**Authors:** Dasom Kim, Jisoo Jo, Jun-Sik Lim, Sukhyun Ryu

**Author notes:** These authors are equally contributed (Dasom Kim, Jisoo Jo).

## Abstract

South Korea is experiencing the community transmission of the SARS-CoV-2 Omicron variant (B.1.1.529). We estimated that the mean (± standard deviation) serial interval was 2.22 (± 1.62) days, and the basic reproduction number was 1.90 (95% Credible Interval, 1.50–2.43) for the Omicron variant outbreak in South Korea.

## Main text

While the SARS-CoV-2 Delta variant is now dominant worldwide, the emergence of the SARS-CoV-2 Omicron variant (B.1.1.529) is causing global concern with its increasing spread.^1^ However, whether the Omicron variant has a faster transmission and/or greater transmissibility in comparison with the Delta variant remains unclear. In the present study, we estimated serial interval and basic reproduction number (*R*_*0*_), involved in the community transmission of the Omicron variant that was initiated from imported cases in South Korea.

We collected publicly available data of the case of the Omicron variant from the Korean public health authorities.^2, 3^ To identify the Omicron variant, public health authorities conducted whole-genome sequencing from respiratory samples from the epidemiologically linked individuals with the omicron variant who tested positive for SARS-CoV-2 by RT-PCR. The Omicron variant case included occurred during November 25, 2021 – December 16, 2021, when the community transmission that was initiated from imported cases reported from the Korea Disease Control and Prevention Agency. The data included contact tracing with other reported cases of the Omicron variant and demographic characteristics of the cases, including age, date of symptom onset, source of infection.

We computed the serial interval as the number of days between the infector and that of the infectee in the transmission pairs. Furthermore, we estimated the serial interval distribution by fitting a normal distribution.^4^

For the newly emerging infectious diseases, the number of cases increases exponentially in the initial phase of an outbreak. The observed value of the exponential growth rate (*r*) is often related to the basic reproductive number, *R*_*0*_. ^4, 5^

In this epidemic theory, *R*_*0*_ can be defined as

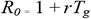

where *T*_*g*_ is the mean generation interval, defined as the mean time interval between the infections of the infector and infectee in a transmission chain. Because the time of infection of the case is usually unobserved, we used the serial interval distribution acquired from this study as a proxy of generation time distribution.^4, 6^

Of the total 131 cases, 56% of cases (n=74) were linked with the church and of these, 31% (n=23) of the cases received two doses of COVID-19 vaccination (Table 1 and Figure 1). We identified 18 transmission pairs having the date of symptom onset for both infector and infectee. The estimated mean serial interval was 2.22 days (95% Credible Interval [CrI], 1.48–2.97) and the standard deviation of the serial interval estimate was 1.62 days (95% CrI, 0.87–2.37) (Figure 2). The *R*_*0*_ in the outbreak was estimated to be 1.90 (95% CrI 1.50–2.43).

**Table 1.**
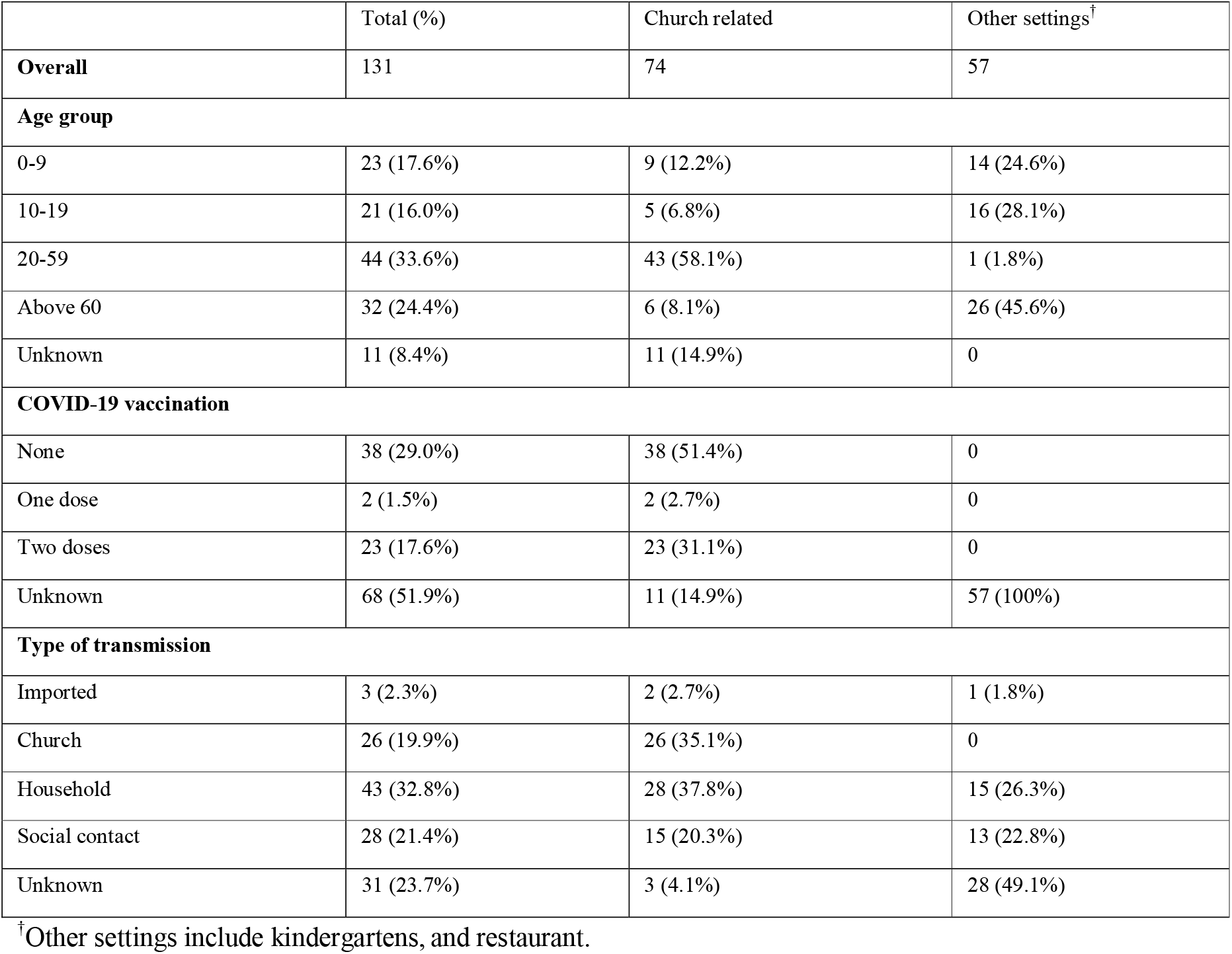
Demographic characteristics of 131 confirmed cases of the SARS-CoV-2 Omicron variant in South Korea.

|  | Total (%) | Church related | Other settings<sup>†</sup> |
| --- | --- | --- | --- |
| **Overall** | 131 | 74 | 57 |
| **Age group** |  |  |  |
| 0-9 | 23 (17.6%) | 9 (12.2%) | 14 (24.6%) |
| 10-19 | 21 (16.0%) | 5 (6.8%) | 16 (28.1%) |
| 20-59 | 44 (33.6%) | 43 (58.1%) | 1 (1.8%) |
| Above 60 | 32 (24.4%) | 6 (8.1%) | 26 (45.6%) |
| Unknown | 11 (8.4%) | 11 (14.9%) | 0 |
| **COVID-19 vaccination** |  |  |  |
| None | 38 (29.0%) | 38 (51.4%) | 0 |
| One dose | 2 (1.5%) | 2 (2.7%) | 0 |
| Two doses | 23 (17.6%) | 23 (31.1%) | 0 |
| Unknown | 68 (51.9%) | 11 (14.9%) | 57 (100%) |
| **Type of transmission** |  |  |  |
| Imported | 3 (2.3%) | 2 (2.7%) | 1 (1.8%) |
| Church | 26 (19.9%) | 26 (35.1%) | 0 |
| Household | 43 (32.8%) | 28 (37.8%) | 15 (26.3%) |
| Social contact | 28 (21.4%) | 15 (20.3%) | 13 (22.8%) |
| Unknown | 31 (23.7%) | 3 (4.1%) | 28 (49.1%) |

<sup>†</sup> Other settings include kindergartens, and restaurant.

**Figure 1.**
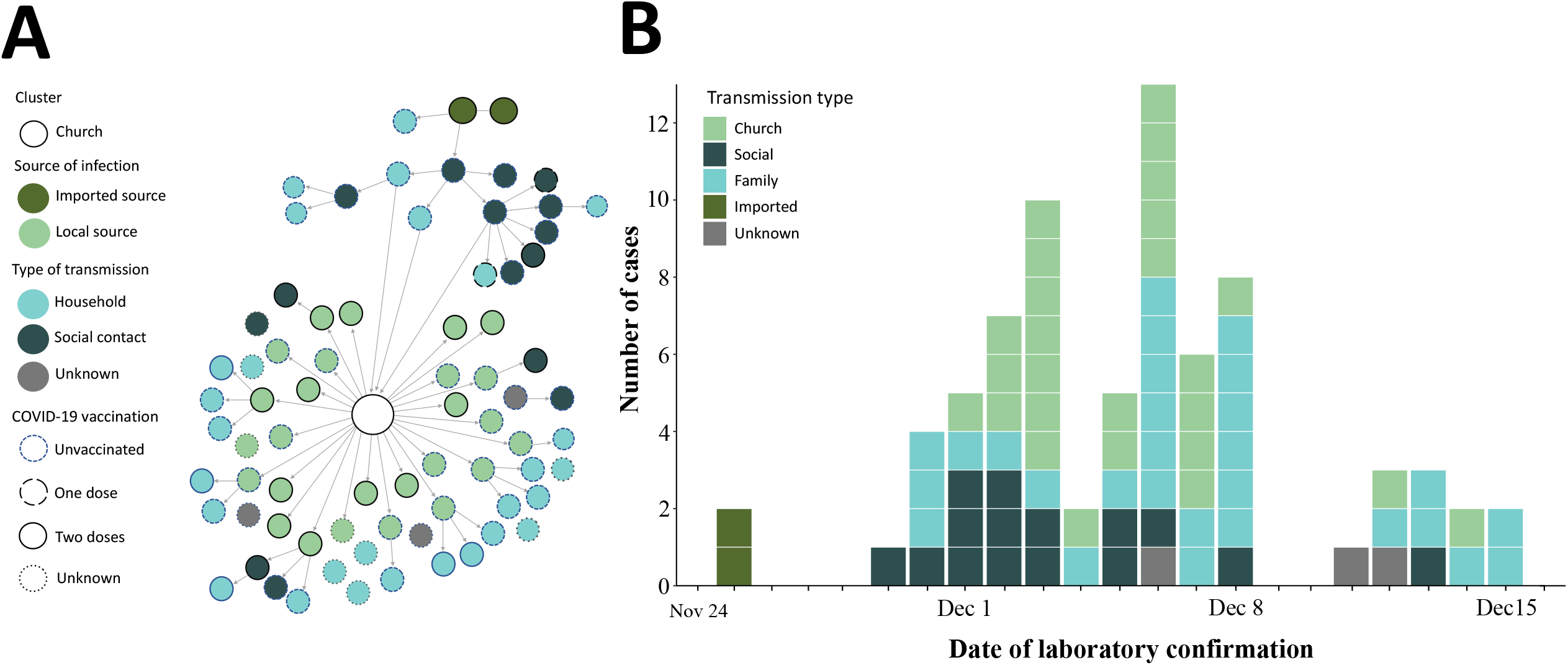
Transmission chain (A) and epidemic curve (B) of laboratory-confirmed SARS-CoV-2 Omicron variant infection associated with the church (n=74) in South Korea.

**Figure 2.**
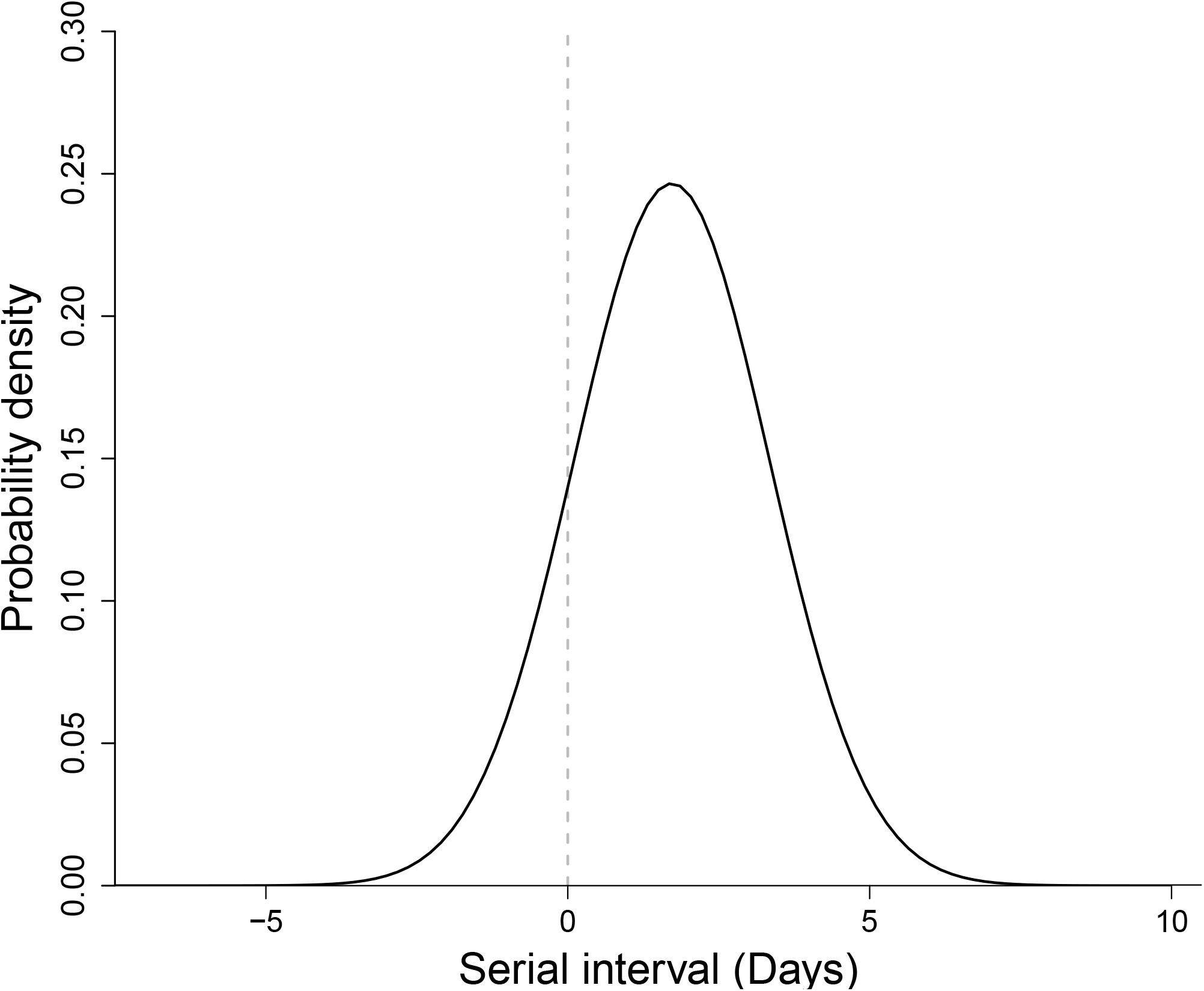
Estimated serial interval distribution of the SARS-CoV-2 Omicron variant. The solid line indicates a fitted normal distribution

Compared with the previous finding of a Korean study on the Delta variant (mean serial interval as 3.3 days and *R*_*0*_ as 1.0),^4^ our findings suggest that the Omicron variant is highly transmissible and likely possesses faster spreading potential in the community. Therefore, quarantine, rapid contact tracing, isolation of asymptomatic contacts, strict adherence to public health measures, and COVID-19 vaccination (including booster doses) remain essential to reduce the community transmission of the Omicron variant.

The limitation of the study is that we estimated the serial interval based on symptomatic cases which may overestimate the result. Second, due to the limitation of the data, we have not analyzed the impact of COVID-19 vaccination against transmission. Third, in the early phase of this outbreak, 14-day isolation had been implemented for all individuals who had any risk of exposure to the case. Furthermore, 80% of the Korean population has received two doses of COVID-19 vaccination as of early December 2021. Therefore, our estimate of transmissibility would not be generalizable in resource-limited countries. Additional study in different settings is warranted to provide evidence for the efficient policy decision-making for the public health measures against the Omicron variant transmission.

## Data Availability

All data produced in the present study are available upon reasonable request to the authors

## Acknowledgments

We appreciate the South Korean public health authorities’ response to COVID-19.

## Ethical approval and consent to participate

This study did not require institutional review board approval or informed consent, because all data used were anonymous and publicly available on public health agency websites.

